# Knowledge, Attitude and Practice (KAP) Survey about hepatitis B (HBV) and C (HCV) among migrant populations from Sub-Saharan Africa

**DOI:** 10.1101/2022.11.29.22282852

**Authors:** Frhat M. A. Saaed, Jerry E. Ongerth, Muna H. Ali

## Abstract

**Background:** Hepatitis B and C virus infections are serious blood borne diseases with global health concern particularly in developing countries. The association between survey participant’s knowledge, attitude, and practices (KAP) about HBV and HCV infection is rarely studied, particularly among migrants seeking resettlement. Libya is a main transient station for migrants northward toward Europe and the flow is increasing into Al Kufra from neighboring countries that have endemic intermediate and high HBV and HCV prevalence. The purpose of this exploratory descriptive study in Al Kufra was to assess the level of participating migrant’s KAP, regarding cause, transmission, prevention, and treatment of viral hepatitis due to HBV and HCV.

**Method:** A cross-sectional study was conducted among 674 sub-Saharan African migrants in Al Kufra, Libya from January to October 2021. Migrant’s KAP about HBV and HCV infection was assessed by interview questionnaire. Statistical and data analysis used SPSS version of 25.

**Results:** Out of 700 questionnaires distributed, 674 were returned with a response rate of 96.3%. The study population included 580 (86.1%) males, mean age of 29.6 ± 7.2 SD years. A majority, 508 (75.4%) were illiterate and elementary level, 8 2.483+.232, respectively. Positive significant linear correlations were found between knowledge-attitude (r = 0.524, p < 0.01) knowledge-practice (r = 0.123, p < 0.01) and attitude-practice (r = 0.278, p < 0.01). Ethnicity and education variables were significantly associated with mean KAP. Gender identity and marital status were both significantly associated with mean knowledge and attitude.

**Conclusion:** The migrant population from the neighboring and sub-Saharan African countries have inadequate KAP about HBV and HCV to limit infection.

## Introduction

Viral hepatitis B and C are blood borne viruses that infect millions of people worldwide. Infections with hepatitis B virus (HBV) and C (HCV) virus are leading causes of chronic liver disease, cirrhosis, liver cancer and associated morbidity and mortality worldwide and are a primary indication for liver transplantation [19]. Chronic HBV and HCV infections are responsible for nearly 57% of liver cirrhosis cases (HBV 30%, HCV 27%) and 78% of hepatocellular carcinomas (HBV 53%, HCV 25%), the third most common cause of cancer deaths worldwide [12, 24, 25].

The endemicity of HBV is high in developing areas, including Africa, the Middle East and Southeast Asia, where at least 8% of the population are HBV chronic carriers [39, 24]. The largest burden of morbidity and mortality from chronic liver disease continues to be in nations of the developing world. For example, 5–10% of the adult population in East Asia and sub-Saharan Africa are estimated to have chronic HBV infection [28]. The highest reported prevalence of chronic HCV is in Africa. Egypt has the highest prevalence of HCV, with some studies reporting HCV antibody positive rates of up to 15%, with an estimated 10% with chronic viremia [14, 11]. Viral hepatitis disproportionately affects sub-Saharan Africans populations [10, 13, 31, 34, 41, 42] with high rates of migration from countries where childhood HBV vaccination has only recently been implemented [15].

The hepatitis B vaccine is the mainstay of HBV prevention [38]. No vaccine is available for HCV, dictating the importance of education and precautions [38]. These viruses are blood- and bodily fluid-borne, transmitted most commonly through sharing of injection equipment, reuse or poorly sterilized of medical equipment, especially syringes and needles and sharp objects, sexual contact, and mother to child transmission [4, 16, 21].

Countries of high HBV & HCV prevalence are often economically disadvantaged and areas of conflict that are a source of migrants seeking stable high-income regions including Europe and North America. This poses challenges to the public health and immigration systems in the host nations [28] and countries along migration routes [26]. In 2020 the total of international migrants was 280.6 million, making this group the 5th most populous nation [38; 22]. Many migrants move into and remain part of ethnic minority groups maintaining traditional social and cultural behaviors that are reinforced and persist and may have adverse implications for exposure to HBV and HCV [30].

Countries such as Libya along migration routes from countries with high prevalence of viral hepatitis, e.g., sub-Saharan African countries have special problems. Migrants typically suffer from infectious diseases that are more prevalent in countries of origin [14, 26]. Migrants from neighboring and sub-Saharan African countries have been cited as one of the major factors spreading the chronic liver disease and hepatocellular carcinoma with risks of disease transmission to the local population [8, 29]. Lack of access to health care services, lack of basic knowledge, poor to nonexistent personal hygiene, and the inability to obtain information about the transmission and prevention all contribute to the importance of defining issues leading to improvement.

The knowledge, attitudes, and practices (KAP) about viral hepatitis transmission, prevention, and liver disease progression among immigrant populations from the neighboring and sub-Saharan African countries is likely to have significant bearing on transmission. Studies show that awareness of chronic HBV or HCV in most populations is low but that among migrant populations it tends to be even lower [30; 36]. A combination of factors contributes to disease propagation, including poor knowledge of the diseases, their risk factors and symptoms, lack of access to healthcare and health information, stigma associated with disease, as well as the lack of symptoms from the early stages of liver disease such as viral hepatitis [30; 33; 34]. General knowledge in immigrant populations is not adequate to avoid blood borne diseases. Social and cultural norms, unsafe sexual intercourse, exposure to the blood or body fluids of an infected person, and intravenous drug use may increase the risk of HBV and HCV infection. Furthermore, minimal education and resources, isolation, and stress contribute the refugees tendency to engage in risky behavior, increasing the risk of these infections [28]. The purpose of this study is to determine the state of knowledge, attitudes, and practices related to HBV and HCV, transmission, consequences, treatment and prevention.

## Method

The study was conducted from January 1 to October 31, 2021 at the Libyan Red Crescent, Al Kufra branch laboratory in a high migrant area. The study area is located in the southeast of Libya having borders with Egypt to the East and with Sudan and Chad to the South. It is 1700 km from Tripoli, the capital city of Libya and 900 km South of the nearest Mediterranean port. The study population of migrants attending at the Libyan Red Crescent Al Kufra branch laboratory included 674 apparently healthy males and females, aged 21–52 years. Knowledge, attitudes, and practices related to HBV and HCV infection were assessed by questionnaire specially designed to permit literacy-independent answers. Questions concerned awareness, beliefs, and attitudes focusing on the causes, transmission, prevention, and treatment of viral hepatitis (Appendix 1-3). Subject categories included behaviors contributing to infection risk with both HBV and HCV; sociodemographic data; and practices of the study population regarding viral hepatitis. A pre-designed, structured questionnaire sheet was completed inside the laboratory by direct personal interview with each migrant individually. The questionnaire was developed and adapted through extensive literature review in English language [1; 5; 20; 31; 40]. The questions in knowledge section were further divided into 3 main sections (see Appendix A): the first 19 knowledge questions were general on viral hepatitis diseases; seven significant viral hepatitis transmission risk factors were included in question 20. Question 21 includes eight common complications of HBV and HCV infections. The attitude and practice section of questionnaire consist of 13 and 12 questions, respectively. The demographic items included age, gender, education level, and ethnicity.

## Statistical Analysis

Data were analyzed using Statistical Package for Social Sciences (SPSS) v. 25.0. The relationship between independent categorical variables and the main outcomes of the study (knowledge, attitude, and practice related to transmission and prevention of HBV and HCV infection) were tested using Chi-square test. The p-value < 0.05 was considered to be significant. Descriptive statistics were used to illustrate migrants’ demographic characteristics. Categorical variables were measured as percentages while continuous variables were expressed as mean ± standard deviation. Normality of data was tested using Kolmogorov-Smirnov test. Inferential statistics involving Chi-square test, Mann-Whitney U test, Kruskal Wallis H test were used to assess the difference while Spearman’s rank correlation coefficient was used to evaluate the relationship between the study variables. Cronbach’s alpha was calculated as an indication of internal consistency.

## Ethics Approval

Written informed consent was obtained from the study participants after they have been clearly briefed about the objective of the study. They were informed about their right to refuse to participate in the study.

## Results

Descriptive statistics for each item in the questionnaire are given in Appendix Tables A1, A2, and A3. A total of 674 African migrants from sub-Saharan Africa, 43.8% Eritrean, 26.6%, Somalian, 21.7%, Ethiopian, and 8% Sudanese, participated in this study (Table 1). Among the 674 participants, 87.5% were married, 86.1% were male and 13.9% female. The age range was 21–52 years old with a mean age of 29.6 years. A majority of the participants, 75.4%, were illiterate or had a basic education level, while 24.6% of participants had a high school level or above (Table 1).

**Table 1.**
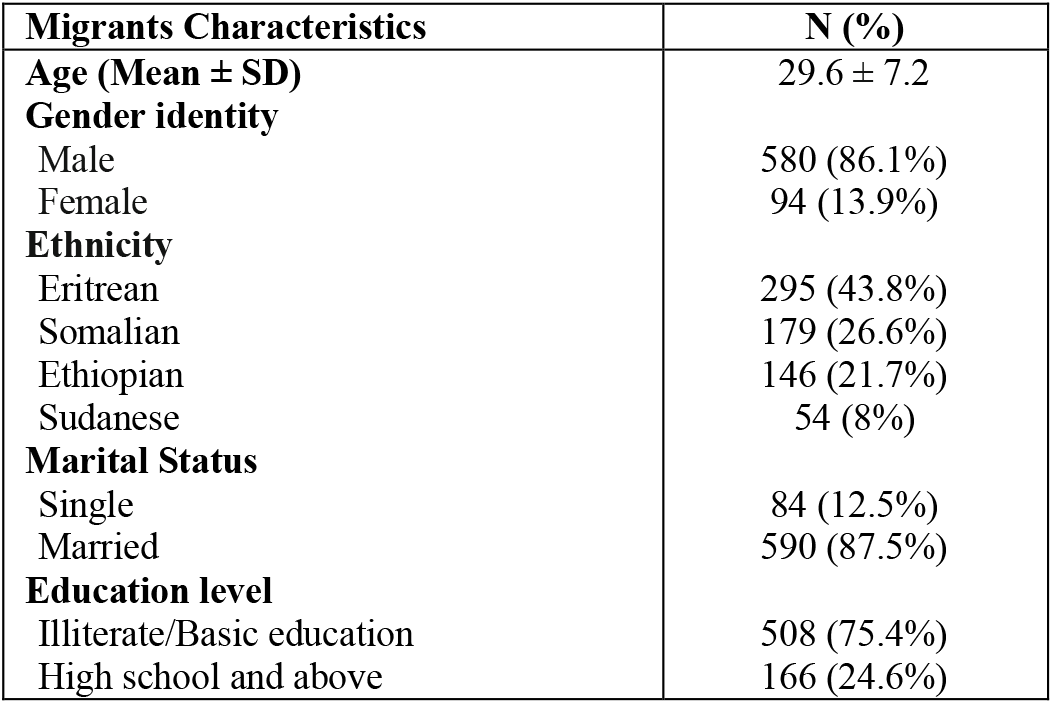
Sub-Saharan African migrant study population demography (n=674)

Of the 700 distributed questionnaires, 674 were returned with a response rate of 96.3%. The Kolmogorov-Smirnov test revealed non normal distribution of the data. The Cronbach’s alpha of the questionnaire was 0.76 for individual domains, maximum Cronbach’s alpha was for attitude section followed by practices and knowledge section. The reason for low Cronbach’s alpha for knowledge domain might be due to the number of questions as compared to attitude and practices sections.

Overall, knowledge regarding HBV and HCV was poor, Table 1a. more than 40% of participants answered Don’t know to the 37 questions reflecting understanding. Most (34 of 37) questions had an answer either correct or not, of these, only 16% of migrants’ answers were correct. For questions of overall indicative knowledge, more than 8 out of 10 migrants, 83.0%, had never heard of viral hepatitis. Less than 1 out of 10 migrants, 7.9%, could identify hepatitis B and C as viral infections (Table 2a). More than 90% of participants had no knowledge about the transmission of HBV and HCV and did not know that a simple test could identify HBV and HCV infections.

**Table 2a.**
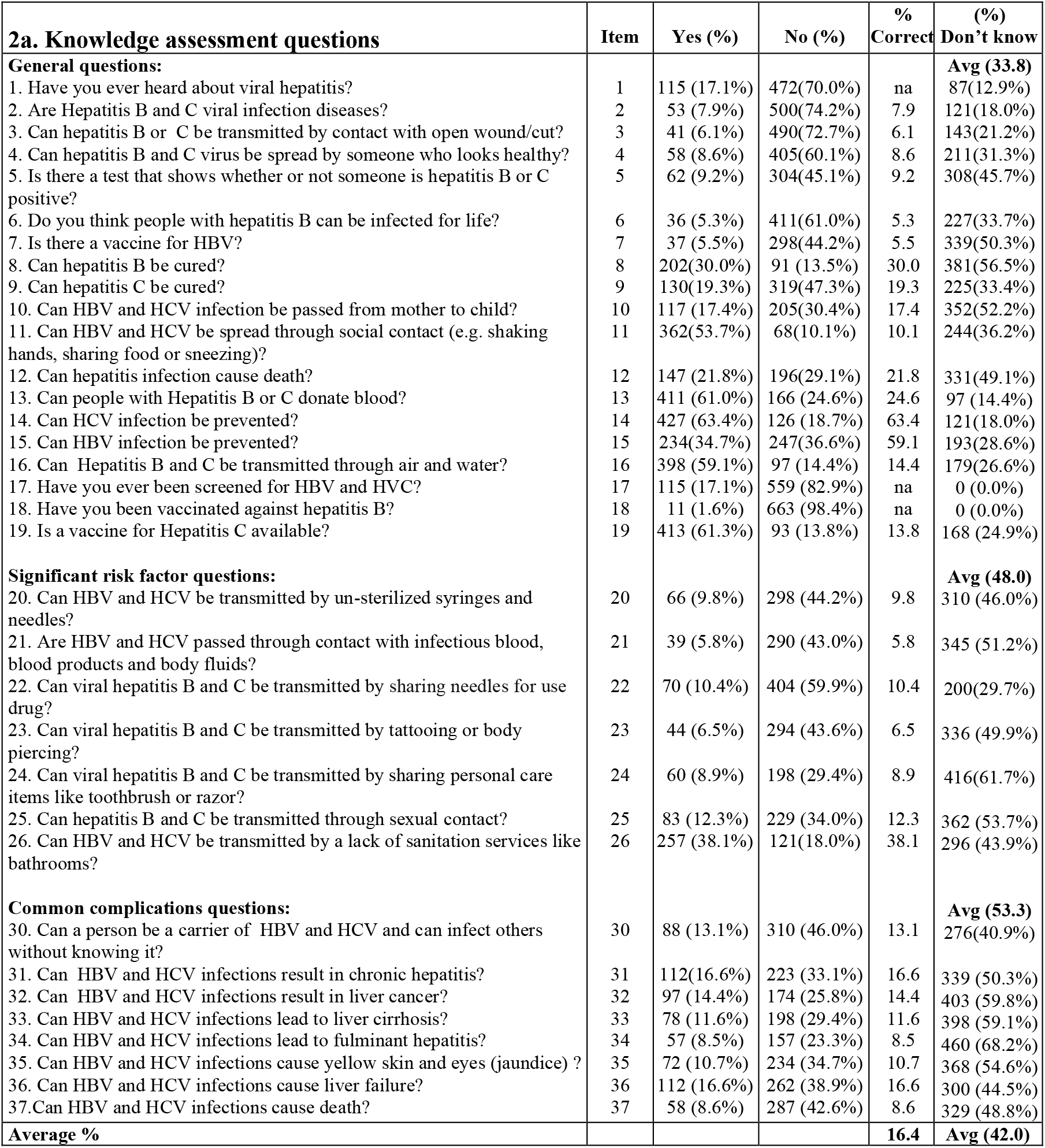
Knowledge segment, KAP study of migrant African populations regarding HBV and HCV infection in Al Kufra, Libya, 2021 (n = 674; 609 male (M), 94 female (F))

| **2a. Knowledge assessment questions** | **Item** | **Yes (%)** | **No (%)** | **% Correct** | **(%) Don't know** |
| --- | --- | --- | --- | --- | --- |
| **General questions:** |  |  |  |  | **Avg (33.8)** |
| 1. Have you ever heard about viral hepatitis? | 1 | 115 (17.1%) | 472(70.0%) | na | 87(12.9%) |
| 2. Are Hepatitis B and C viral infection diseases? | 2 | 53 (7.9%) | 500(74.2%) | 7.9 | 121(18.0%) |
| 3. Can hepatitis B or C be transmitted by contact with open wound/cut? | 3 | 41 (6.1%) | 490(72.7%) | 6.1 | 143(21.2%) |
| 4. Can hepatitis B and C virus be spread by someone who looks healthy? | 4 | 58 (8.6%) | 405(60.1%) | 8.6 | 211(31.3%) |
| 5. Is there a test that shows whether or not someone is hepatitis B or C positive? | 5 | 62 (9.2%) | 304(45.1%) | 9.2 | 308(45.7%) |
| 6. Do you think people with hepatitis B can be infected for life? | 6 | 36 (5.3%) | 411(61.0%) | 5.3 | 227(33.7%) |
| 7. Is there a vaccine for HBV? | 7 | 37 (5.5%) | 298(44.2%) | 5.5 | 339(50.3%) |
| 8. Can hepatitis B be cured? | 8 | 202(30.0%) | 91 (13.5%) | 30.0 | 381(56.5%) |
| 9. Can hepatitis C be cured? | 9 | 130(19.3%) | 319(47.3%) | 19.3 | 225(33.4%) |
| 10. Can HBV and HCV infection be passed from mother to child? | 10 | 117 (17.4%) | 205(30.4%) | 17.4 | 352(52.2%) |
| 11. Can HBV and HCV be spread through social contact (e.g. shaking hands, sharing food or sneezing)? | 11 | 362(53.7%) | 68(10.1%) | 10.1 | 244(36.2%) |
| 12. Can hepatitis infection cause death? | 12 | 147 (21.8%) | 196(29.1%) | 21.8 | 331(49.1%) |
| 13. Can people with Hepatitis B or C donate blood? | 13 | 411 (61.0%) | 166 (24.6%) | 24.6 | 97 (14.4%) |
| 14. Can HCV infection be prevented? | 14 | 427 (63.4%) | 126 (18.7%) | 63.4 | 121(18.0%) |
| 15. Can HBV infection be prevented? | 15 | 234(34.7%) | 247(36.6%) | 59.1 | 193(28.6%) |
| 16. Can Hepatitis B and C be transmitted through air and water? | 16 | 398 (59.1%) | 97 (14.4%) | 14.4 | 179(26.6%) |
| 17. Have you ever been screened for HBV and HVC? | 17 | 115 (17.1%) | 559 (82.9%) | na | 0 (0.0%) |
| 18. Have you been vaccinated against hepatitis B? | 18 | 11 (1.6%) | 663 (98.4%) | na | 0 (0.0%) |
| 19. Is a vaccine for Hepatitis C available? | 19 | 413 (61.3%) | 93 (13.8%) | 13.8 | 168 (24.9%) |
| **Significant risk factor questions:** |  |  |  |  | **Avg (48.0)** |
| 20. Can HBV and HCV be transmitted by un-sterilized syringes and needles? | 20 | 66 (9.8%) | 298 (44.2%) | 9.8 | 310 (46.0%) |
| 21. Are HBV and HCV passed through contact with infectious blood, blood products and body fluids? | 21 | 39 (5.8%) | 290 (43.0%) | 5.8 | 345 (51.2%) |
| 22. Can viral hepatitis B and C be transmitted by sharing needles for use drug? | 22 | 70 (10.4%) | 404 (59.9%) | 10.4 | 200(29.7%) |
| 23. Can viral hepatitis B and C be transmitted by tattooing or body piercing? | 23 | 44 (6.5%) | 294 (43.6%) | 6.5 | 336 (49.9%) |
| 24. Can viral hepatitis B and C be transmitted by sharing personal care items like toothbrush or razor? | 24 | 60 (8.9%) | 198 (29.4%) | 8.9 | 416(61.7%) |
| 25. Can hepatitis B and C be transmitted through sexual contact? | 25 | 83 (12.3%) | 229 (34.0%) | 12.3 | 362 (53.7%) |
| 26. Can HBV and HCV be transmitted by a lack of sanitation services like bathrooms? | 26 | 257 (38.1%) | 121(18.0%) | 38.1 | 296 (43.9%) |
| **Common complications questions:** |  |  |  |  | **Avg (53.3)** |
| 30. Can a person be a carrier of HBV and HCV and can infect others without knowing it? | 30 | 88 (13.1%) | 310 (46.0%) | 13.1 | 276(40.9%) |
| 31. Can HBV and HCV infections result in chronic hepatitis? | 31 | 112(16.6%) | 223 (33.1%) | 16.6 | 339 (50.3%) |
| 32. Can HBV and HCV infections result in liver cancer? | 32 | 97 (14.4%) | 174 (25.8%) | 14.4 | 403 (59.8%) |
| 33. Can HBV and HCV infections lead to liver cirrhosis? | 33 | 78 (11.6%) | 198 (29.4%) | 11.6 | 398 (59.1%) |
| 34. Can HBV and HCV infections lead to fulminant hepatitis? | 34 | 57 (8.5%) | 157 (23.3%) | 8.5 | 460 (68.2%) |
| 35. Can HBV and HCV infections cause yellow skin and eyes (jaundice) ? | 35 | 72 (10.7%) | 234 (34.7%) | 10.7 | 368 (54.6%) |
| 36. Can HBV and HCV infections cause liver failure? | 36 | 112 (16.6%) | 262 (38.9%) | 16.6 | 300 (44.5%) |
| 37. Can HBV and HCV infections cause death? | 37 | 58 (8.6%) | 287 (42.6%) | 8.6 | 329 (48.8%) |
| **Average %** |  |  |  | **16.4** | **Avg (42.0)** |

**Table 2b.** Attitude assessment of the migrant population regarding HBV and HCV.

| **2b. Attitude assessment questions** | **Item no.** | **Definitely Agree** | **Moderately agree** | **Neutral** | **Moderately disagree** |
| --- | --- | --- | --- | --- | --- |
| 1. I have no concern for being infected with hepatitis B or C. | 1 | 83(12.3%) | 164(24.3%) | 295(43.8%) | 80(11.9%) |
| 2. Hepatitis B vaccine is safe and effective. | 2 | 23(3.4%) | 36(5.3%) | 369(54.7%) | 176(26.1%) |
| 3. All African migrant populations should be tested for hepatitis B or C before they receive health care. | 3 | 38(5.6%) | 74(11.0%) | 259(38.4%) | 198(29.4%) |
| 4. I don't feel comfortable living with people with hepatitis B or C. | 4 | 20(3.0%) | 64(9.5%) | 446(66.2%) | 121(18.0%) |
| 5. I believe that hepatitis infections can be transmitted through food. | 5 | 49(7.3%) | 182(27.0%) | 340(50.4%) | 78(11.6%) |
| 6. I think hepatitis B is a curable disease. | 6 | 87(12.9%) | 167(24.8%) | 369(54.7%) | 27(4.0%) |
| 7. I think that hepatitis B and C is a serious public health problem. | 7 | 119(17.7%) | 138(20.5%) | 285(42.3%) | 96(14.2%) |
| 8. I can get hepatitis infection. | 8 | 45(6.7%) | 91(13.5%) | 341(50.6%) | 138(20.5%) |
| 9. Viral hepatitis is something to worry about. | 9 | 33(4.9%) | 74(11.0%) | 260(38.6%) | 221(32.8%) |
| 10. I believe that vaccination could prevent transmission. | 10 | 49(7.3%) | 111(16.5%) | 244(36.2%) | 197(29.2%) |
| 11. I believe that healthy people need vaccination. | 11 | 27(4.0%) | 117(17.4%) | 221(32.8%) | 243(36.1%) |
| 12. I believe that you need a vaccination at your age. | 12 | 22(3.3%) | 70(10.4%) | 503(74.6%) | 41(6.1%) |
| 13. I believe that vaccine can be free or low cost through certain programs. | 13 | 34(5.0%) | 38(5.6%) | 267(39.6%) | 188(27.9%) |

**Table 2c.**
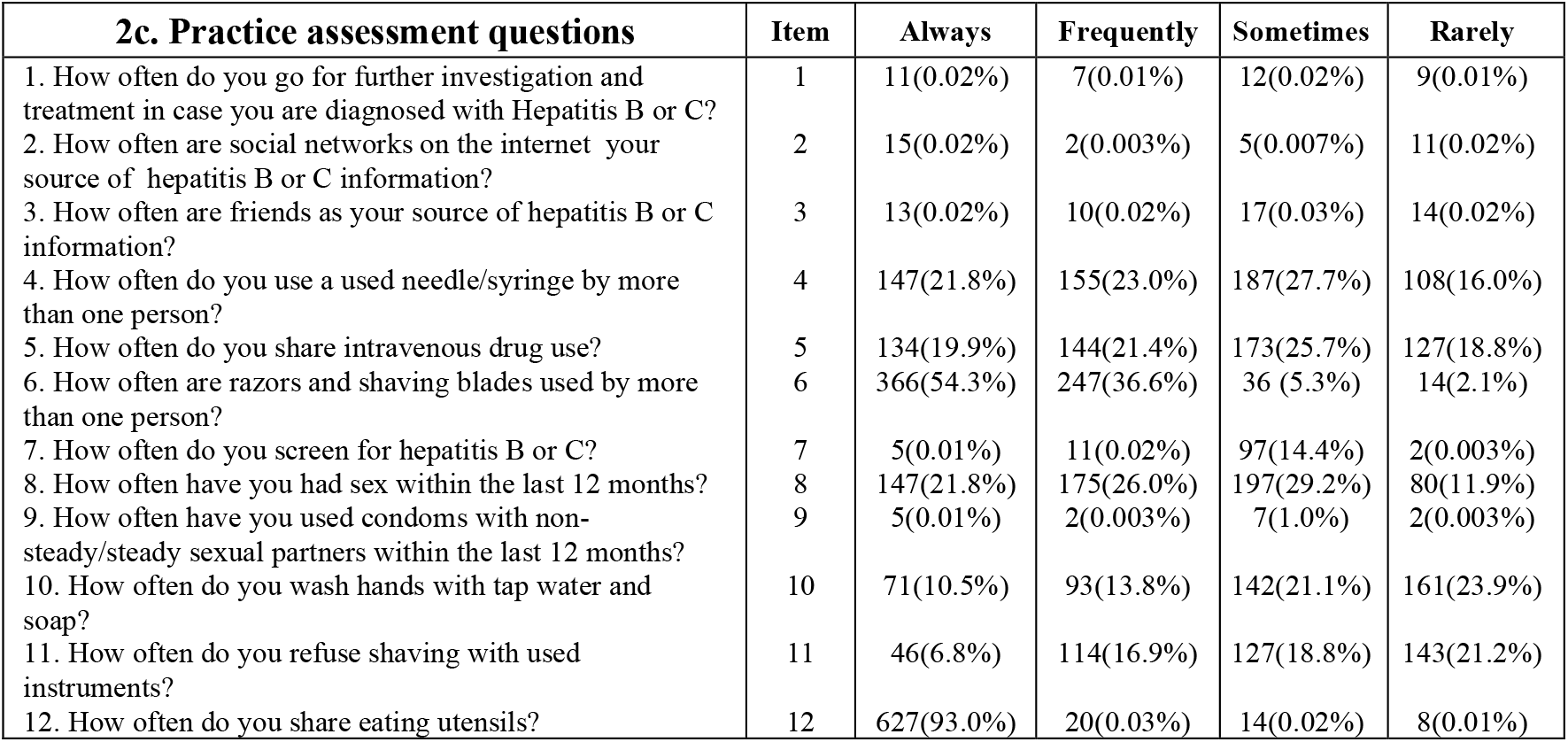
Practice assessment of migrant the population regarding HBV and HCV.

| 2c. Practice assessment questions | Item | Always | Frequently | Sometimes | Rarely |
| --- | --- | --- | --- | --- | --- |
| 1. How often do you go for further investigation and treatment in case you are diagnosed with Hepatitis B or C? | 1 | 11(0.02%) | 7(0.01%) | 12(0.02%) | 9(0.01%) |
| 2. How often are social networks on the internet your source of hepatitis B or C information? | 2 | 15(0.02%) | 2(0.003%) | 5(0.007%) | 11(0.02%) |
| 3. How often are friends as your source of hepatitis B or C information? | 3 | 13(0.02%) | 10(0.02%) | 17(0.03%) | 14(0.02%) |
| 4. How often do you use a used needle/syringe by more than one person? | 4 | 147(21.8%) | 155(23.0%) | 187(27.7%) | 108(16.0%) |
| 5. How often do you share intravenous drug use? | 5 | 134(19.9%) | 144(21.4%) | 173(25.7%) | 127(18.8%) |
| 6. How often are razors and shaving blades used by more than one person? | 6 | 366(54.3%) | 247(36.6%) | 36 (5.3%) | 14(2.1%) |
| 7. How often do you screen for hepatitis B or C? | 7 | 5(0.01%) | 11(0.02%) | 97(14.4%) | 2(0.003%) |
| 8. How often have you had sex within the last 12 months? | 8 | 147(21.8%) | 175(26.0%) | 197(29.2%) | 80(11.9%) |
| 9. How often have you used condoms with non-steady/steady sexual partners within the last 12 months? | 9 | 5(0.01%) | 2(0.003%) | 7(1.0%) | 2(0.003%) |
| 10. How often do you wash hands with tap water and soap? | 10 | 71(10.5%) | 93(13.8%) | 142(21.1%) | 161(23.9%) |
| 11. How often do you refuse shaving with used instruments? | 11 | 46(6.8%) | 114(16.9%) | 127(18.8%) | 143(21.2%) |
| 12. How often do you share eating utensils? | 12 | 627(93.0%) | 20(0.03%) | 14(0.02%) | 8(0.01%) |

A majority, 86.5% falsely believed that hepatitis B could be cured (or didn’t know), while nearly half, 47.3% believed that hepatitis C is not curable although a third (33.4%) ‘did not know’. The vast majority, 98.4% did not know that an HBV vaccine existed and only 1.6% of the migrants had been vaccinated. Moreover, 82.9% had never been screened for HBV.

Knowledge of risk factors was also very poor. The overwhelming majority had no understanding that HBV and HCV could be transmitted by mechanisms including: by un-sterilized syringes, 90.2%; by blood products and body fluids, 94.2%; and by sharing needles, 89.6%. Furthermore, similarly high majorities did not understand that both HBV & HCV could be transmitted by tattooing, 93.5%, sharing personal care items 91.0%, and through sexual contact, 87.7%, (Table 2). Among the 674 migrant participants, only ca. one in 10 understood that complications of chronic hepatitis include liver cancer, liver cirrhosis, fulminant hepatitis, and liver failure (Table 2a).

Results of attitude assessment indicated that nearly 90% of the migrants were not concerned about being infected with HBV or HCV and did not consider the infections a public health problem Table 2b. Only about 20% believed that they could get HBV or HCV infections and 15% were not worried about it. Only 15% agreed that migrants should be tested and less than 10% of participants agreed that the HBV vaccine is safe and effective (Table 2b). Similar to the knowledge response (Table 1a), less than 10% of participants believed or didn’t know that HBV was not curable. Reflecting lack of concern, more than 90% of participants we felt comfortable living with people with hepatitis B or C, while 34.3% of the migrants believed that HBV and HCV infection could be transmitted by food. More than two thirds of participants showed a negative attitude towards vaccination (Table 2b).

Participating migrant responses to practice assessment questions indicate many unsafe practices that may result in exposure to blood- and body fluid-borne diseases. Very few, 4.3%, indicated that they would seek further diagnosis and treatment if they tested positive for HBV or HCV, Table 2c. Very few indicated a ready source of information about hepatitis B or C, social networks, 4.9%, or friends, 8.0%. Responses reflecting lack of knowledge or reliable source of information, very high proportions of participants indicated high-risk practices for transmission: 84% indicted shared use of injecting materials; 81% indicated shared intravenous drug use; almost all, 98% indicated sharing shaving materials; and with a high rate of sexual activity, ca. 90%, almost none indicated use of condoms. Reflecting lack of understanding the implications of high-risk practices, very few, ca 15%, ever went for hepatitis B or C screening.

Inferential statistics, i.e., Mann-Whitney U and Kruskal Wallis H tests were applied to compare scores of each domain with various demographic factors Table 3. Mean knowledge and attitude scores of male and female differed for p<0.05. Male showed higher knowledge and attitude than females. Eritrean migrants had significantly higher mean knowledge and practice scores than other demographic groups. The mean knowledge scores were significantly higher among single than married migrants, while the mean practice score of married migrants was significantly higher than among single migrants. Finally, significantly higher differences were observed between mean knowledge and attitude scores for migrants with high school education compared to those with illiterate/basic education. However, those having high school education had significantly lower mean practice scores.

**Table 3.** Comparison of demographic characteristics for mean knowledge, attitude and practice scores.

| Demographic variables | K.score<br>0.794+.204<br>Mean rank | P-value | A.score<br>2.906+.380<br>Mean rank | P-value | P.score<br>2.483+.232<br>Mean rank | P-value |
| --- | --- | --- | --- | --- | --- | --- |
| **Gender identity\*\*** |  |  |  |  |  |  |
| Male | 355.27 | 0.001 | 343.47 | 0.047 | 335.08 | 0.418 |
| Female | 227.85 |  | 300.66 |  | 352.43 |  |
| **Ethnicity\*** |  |  |  |  |  |  |
| Eritrean | 362.56 | 0.024 | 371.98 | 0.001 | 363.24 | 0.007 |
| Somalian | 320.64 |  | 288.41 |  | 317.97 |  |
| Ethiopian | 309.08 |  | 305.26 |  | 331.08 |  |
| Sudanese | 333.32 |  | 399.03 |  | 278.99 |  |
| **Marital Status\*\*** |  |  |  |  |  |  |
| Single | 379.88 | 0.033 | 340.07 | 0.897 | 292.05 | 0.021 |
| Married | 331.47 |  | 337.13 |  | 343.97 |  |
| **Education level\*\*** |  |  |  |  |  |  |
| Illiterate/Basic education | 291.61 | 0.001 | 305.40 | 0.001 | 352.02 | 0.001 |
| High school and above | 477.94 |  | 435.72 |  | 293.06 |  |

\* Kruskal Wallis Test, \*\* Mann Whitney Test, *P-value < 0.05 is statistically significant.*

A correlation between different domains of questionnaire was also assessed. The correlation revealed significant positive linear correlations between knowledge-attitude (r = 0.524, p < 0.01) knowledge-practice (r = 0.123, p < 0.01) and attitude-practice (r = 0.278, p < 0.01), as shown in Table 4.

**Table 4.** Correlation between knowledge, attitude, and practice scores.

| Variables | Correlation coefficient | P-value* |
| --- | --- | --- |
| Knowledge, attitude | 0.524 | 0.01 |
| Knowledge, practice | 0.123 | 0.01 |
| Practice, Attitude | 0.278 | 0.01 |

\*Correlation significant at 0.01 level (2 tailed).

Using chi-square test, for knowledge scores of the study participants were divided into two categories: good, and poor. Attitude and practice scores of the study participants were divided into three categories: good, fair and poor. On this basis, knowledge was ‘Poor’, >85%, across all categories of gender, ethnicity, and marital status (Table 5). Attitudes could be considered ‘Fair’ for most categories. But practices were considered ‘Poor’ for > 2/3’s of participants across all categories.

**Table 5.**
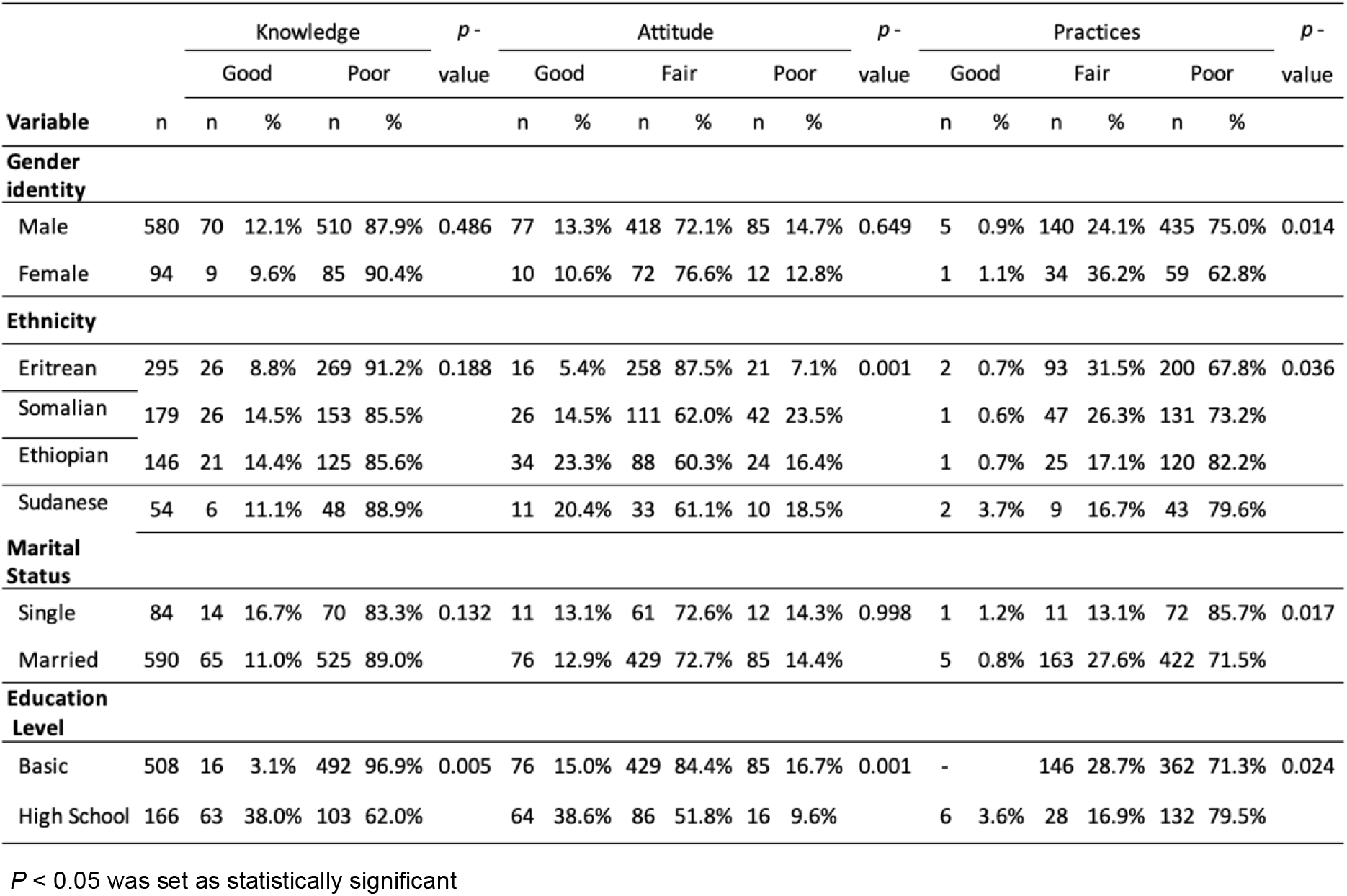
Categorization of the study African migrant populations score on KAP domains.

## Discussion

Currently, the flow of immigrants into Libya from neighboring countries is increasing. However, there are no post-arrival screening mechanisms for hepatitis B and C viruses to assess migrant conditions or to support public health policy or actions. Accordingly, the purpose of this study was to assess the level of participating migrant’s knowledge, attitude, and practices, regarding cause, transmission, prevention and treatment of viral hepatitis due to HBV and HCV. Libya public health professionals are concerned as the country as a whole is classified among geographical regions with low-intermediate endemicity for hepatitis B virus infection and an area of low endemicity for hepatitis C. The prevalence of hepatitis B surface antigen (HBsAg) and anti-HCV antibodies in sub-Saharan African migrants has been found to exceed 30-50% indicating significant cause for concern (27). A plausible reason for this high prevalence appears related to the lack of knowledge about HBV and HCV, their health significance, their means of transmission, of actions for preventing infection.

The present study assessed the participant’s knowledge, attitude, and practices about HBV and HCV infection. Results clearly show poor KAP towards HBV and HCV among migrants at Al Kufra. An overwhelming lack of knowledge about HBV and HCV, their mode of transmission, and the possibility of prevention by vaccination. was observed among participating migrants Most of these sub-Saharan participants did not know typical ways of transmission, prevention, and development of liver disease. In addition, participants were not aware of the different types of viral hepatitis and did not have access to information about their consequences. Only just over 15% the migrants in our study had basic knowledge about HBV and HCV as a disease of liver, consistent with a previous report of an Ethiopia study in which just 11.0% of the refugees know the relationship between liver cancer and hepatitis B and C. Ethiopia has a population-wide prevalence of 7.4 and 3.1% for HBV and HCV respectively [5].

Most of the migrants in this study, 84.12%, did not consider HBV and HCV as a real risk for themselves and for their family. Although this may explain their inadequate practices it almost certainly contributes to high rates of HBV and HCV infection [27]. Migrants participating in this study showed poorly supported attitudes and exceedingly poor practices towards HBV and HCV reflecting their very low state of knowledge important to minimizing risk and infection. A wide range of unsafe practices are evident in the survey responses, exposing the migrants to infection by blood borne diseases.

## Conclusions

In this screening study, the level of knowledge and awareness of migrant African populations transiting Libya was poor about the concepts of infectious risk associated HBV and HCV. Poor knowledge and lack of awareness of the African immigrants about HBV and HCV must contribute to the prevalence of these infections in developing countries compared with developed ones. Survey results demonstrate that knowledge on transmission is lacking, and awareness about HBV and HCV transmission to others is only fair. Lacking information about transmission and prevention migrants are at risk of contracting HBV and HCV emphasizing the need for education to limit the spread of the blood borne disease. Opportunities exist for attention by Libyan health and immigration authorities to assist migrants seeking improved conditions. Poor knowledge and lack of awareness about HBV and HCV among sub-Saharan populations undoubtedly contributes to the spread of these infections in native countries.

## Data Availability

All data produced in the present work are contained in the manuscript

## Appendix

**Table A1.**
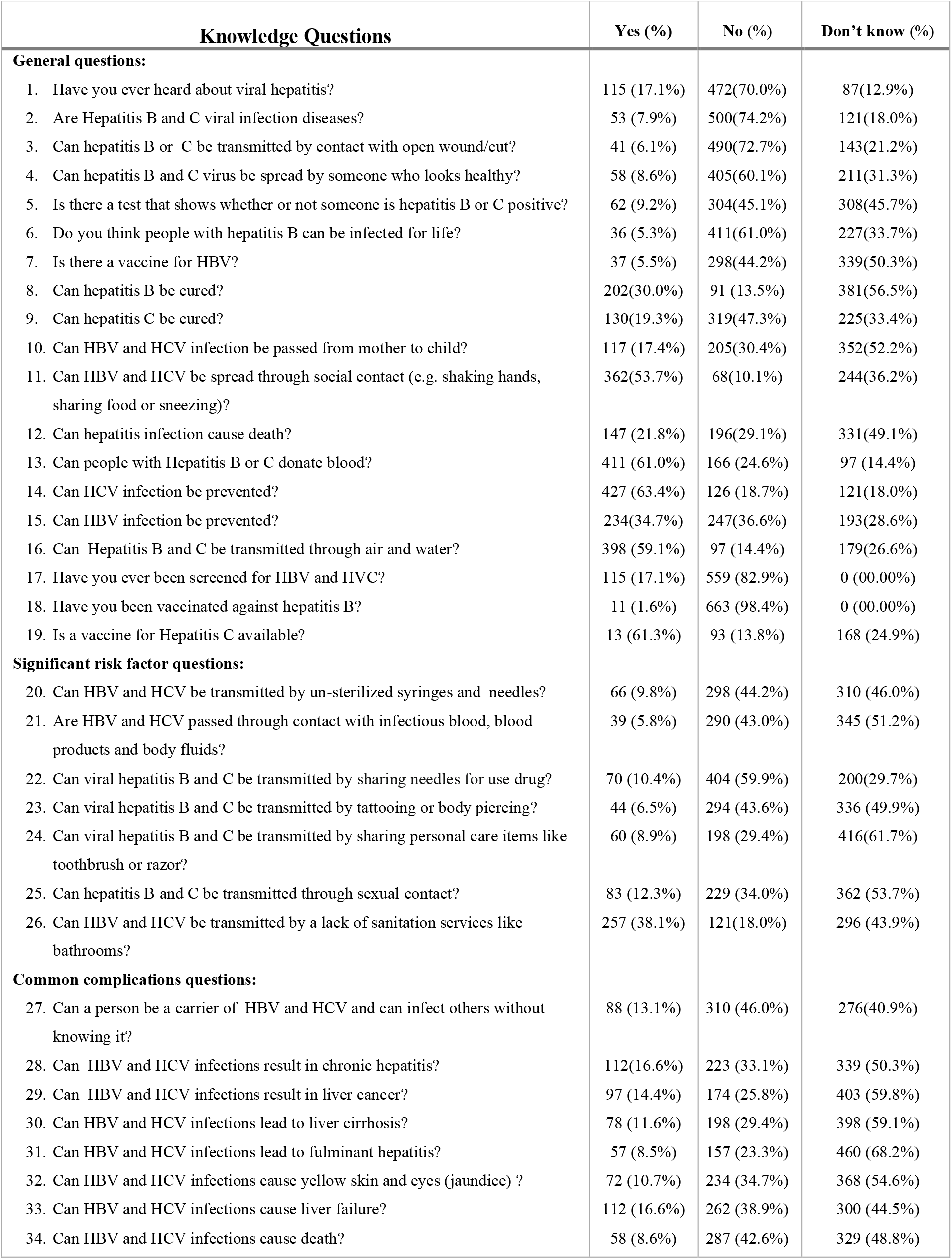
Questions on Knowledge of study participating migrants about HBV and HCV.

**Table A2.** Questions regarding Attitude of participating migrants about HBV and HCV.

| Attitude assessment questions | Definitely Agree | Moderately agree | Neutral | Moderately disagree | Definitely Disagree |
| --- | --- | --- | --- | --- | --- |
| 1. I have no concern for being infected with Hepatitis B or C. | 83(12.3%) | 164(24.3%) | 295(43.8%) | 80(11.9%) | 52(7.7%) |
| 2. Hepatitis B vaccine is safe and effective. | 23(3.4%) | 36(5.3%) | 369(54.7%) | 176(26.1%) | 70(10.4%) |
| 3. All African migrant populations should be tested for hepatitis B or C before they receive health care. | 38(5.6%) | 74(11.0%) | 259(38.4%) | 198(29.4%) | 105(15.6%) |
| 4. I don't feel comfortable to live with people with Hepatitis B or C. | 20(3.0%) | 64(9.5%) | 446(66.2%) | 121(18.0%) | 23(3.4%) |
| 5. I believe that hepatitis infections can be transmitted through food. | 49(7.3%) | 182(27.0%) | 340(50.4%) | 78(11.6%) | 25(3.7%) |
| 6. I think hepatitis B is a curable disease. |  |  |  |  |  |
| 7. I think that hepatitis B and C is a serious public health problem. | 87(12.9%) | 167(24.8%) | 369(54.7%) | 27(4.0%) | 24(3.6%) |
| 8. I can get hepatitis infection. | 119(17.7%) | 138(20.5%) | 285(42.3%) | 96(14.2%) | 36(5.3%) |
| 9. Viral hepatitis is something to worry about. | 45(6.7%) | 91(13.5%) | 341(50.6%) | 138(20.5%) | 59(8.8%) |
| 10. I believe that vaccination could prevent transmission. | 33(4.9%) | 74(11.0%) | 260(38.6%) | 221(32.8%) | 86(12.8%) |
| 11. I believe that healthy people need vaccination. | 49(7.3%) | 111(16.5%) | 244(36.2%) | 197(29.2%) | 73(10.8%) |
| 12. I believe that you need a vaccination at your age. | 27(4.0%) | 117(17.4%) | 221(32.8%) | 243(36.1%) | 66(9.8%) |
| 13. I believe that vaccine can be free or low cost through certain programs. | 22(3.3%) | 70(10.4%) | 503(74.6%) | 41(6.1%) | 38(5.6%) |
|  | 34(5.0%) | 38(5.6%) | 267(39.6%) | 188(27.9%) | 147(21.8%) |

**Table A3.** Questions regarding Practices of participating migrants about HBV and HCV.

| Practice Assessment Questions | Always | Frequently | Sometimes | Rarely | Never |
| --- | --- | --- | --- | --- | --- |
| 1. How often do you go for further investigation and treatment, if you are diagnosed with hepatitis B or C? | 11(01.63%) | 7(01.03%) | 12(01.78%) | 9(01.33%) | 635(94.21%) |
| 2. How often are social networks on the internet your source of hepatitis B or C information? | 15(02.22%) | 2(00.29%) | 5(00.74%) | 11(01.63%) | 641(95.10%) |
| 3. How often are friends as your source of hepatitis B or C information? | 13(1.9%) | 10(1.5%) | 17(2.5%) | 14(2.1%) | 620(92.0%) |
| 4. How often do you use a used needle/syringe by more than one person? | 47(21.8%) | 155(23.0%) | 187(27.7%) | 108(16.0%) | 77(11.4%) |
| 5. How often do you share intravenous drug use? | 134(19.9%) | 144(21.4%) | 173(25.7%) | 127(18.8%) | 96(14.2%) |
| 6. How often are razors and shaving blades used by more than one person? | 366(54.3%) | 247(36.6%) | 36 (5.3%) | 14(2.1%) | 11(1.6%) |
| 7. How often do you screen for hepatitis B or C? | 5(.7%) | 11(1.6%) | 97(14.4%) | 2(.3%) | 559(82.9%) |
| 8. How often have you had sex within the last 12 months? | 147(21.8%) | 175(26.0%) | 197(29.2%) | 80(11.9%) | 75(11.1%) |
| 9. How often have you used condoms with non-steady/steady sexual partners within the last 12 months? | 5(.7%) | 2(.3%) | 7(1.0%) | 2(.3%) | 658(97.6%) |
| 10. How often do you wash hands with tap water and soap? | 71(10.5%) | 93(13.8%) | 142(21.1%) | 161(23.9%) | 207(30.7%) |
| 11. How often do you refuse shaving with used instruments? | 46(6.8%) | 114(16.9%) | 127(18.8%) | 143(21.2%) | 244(36.2%) |
| 12. How often do you share eating utensils? | 627(93.0%) | 20(3.0%) | 14(2.1%) | 8(1.2%) | 5(.7%) |

